# When evidence is not enough: A qualitative exploration of healthcare workers’ perspectives on expansion of two-way texting (2wT) for post-circumcision follow-up in South Africa

**DOI:** 10.1101/2024.10.22.24315946

**Authors:** Isabella Fabens, Calsile Makhele, Nelson Igaba, Khumbulani Moyo, Felex Ndebele, Jacqueline Pienaar, Geoffrey Setswe, Caryl Feldacker

## Abstract

As per national guidelines, in-person follow-up visits after voluntary medical male circumcision (VMMC) are required but may be unnecessary. Two-way texting (2wT) engages clients in post-operative care and triages those with complications to in-person review. 2wT-based telehealth was found to be safe, effective, and efficient. In South Africa, to understand provider perspectives on the 2wT approach and potential for expansion, 20 key informant interviews were conducted with management, clinicians, data officials and support staff involved in 2wT scale-up. Interviews were analyzed using rapid qualitative methods and informed by two implementation science frameworks: the Reach, Effectiveness, Adoption, Implementation and Maintenance (RE-AIM) framework and the Pragmatic, Robust, Implementation and Sustainability Model (PRISM). Participants submitted mixed and multi-faceted feedback, including that 2wT improves monitoring and evaluation of clients and clinical outcomes while also reducing follow-up visits. Challenges included duplicative routine and 2wT reporting systems and perceptions that 2wT increased workload. To improve the likelihood of successful 2wT scale-up in routine VMMC settings, respondents suggested: further 2wT sensitization to ensure clinician and support staff buy-in; a dedicated clinician or nurse to manage telehealth clients; improved dashboards to better visualize 2wT client data; mobilizing 2wT champions at facilities to garner support for 2wT as routine care; and, updating VMMC guidelines to support VMMC telehealth. As attendance at follow-up visits may not be as high as reported, implementing 2wT may require more effort but also brings added benefits of client verification and documented follow-up. The transition from research to routine practice is challenging, but use of RE-AIM and PRISM indicate that it is not impossible. As VMMC funding is decreasing, more effort to share the evidence base for 2wT as a safe, cost-effective, high-quality approach for VMMC follow-up is needed to encourage widespread uptake and adoption.

## Introduction

Although voluntary medical male circumcision (VMMC) is a safe procedure with few adverse events (AEs) [1–6], global VMMC guidelines still require clients to attend two post-operative follow-up visits within 14 days to ensure timely identification and treatment for AEs. This presents an undue burden for providers and clients [7, 8] creating potential barriers to continued VMMC expansion in support of HIV prevention efforts. Therefore, in 2018, the International Training and Education Center for Health (I-TECH) at the University of Washington (UW) implemented a two-way texting (2wT) approach to provide SMS-based telehealth in Zimbabwe, ensuring a safe and more efficient option for VMMC follow-up. In 2021, I-TECH partnered with the Aurum Institute, the Centre for HIV-AIDS Prevention Studies (CHAPS), and Right to Care to expand on this work in South Africa, first conducting a randomized controlled trial (RCT) and then an expansion trial. Similar to Zimbabwe results [9, 10], 2wT was found effective to provide safe, lower-cost, follow-up that greatly benefitted VMMC clients who approved of the approach [8, 11–13]. Previous qualitative study among healthcare workers (HCW) in both Zimbabwe [14, 15] and South Africa [13] found 2wT to be useful, acceptable, and highly usable. However, from the HCW implementation perspective, challenges were identified in duplicative VMMC reporting systems that increased workload, the need for specific 2wT personnel, and the need for improved client wound care counseling to reduce concerns that clients were unable to identify complications swiftly.

In this qualitative study on 2wT expansion potential in South Africa, our objective was to complement previous findings by focusing on new opportunities for adaptation and identifying additional potential pitfalls moving 2wT from research to routine VMMC settings with routine VMMC service delivery teams. Although rigorous implementation science (IS) studies of mobile health (mHealth) interventions remain rare, we used an IS approach to help understand how, why or for whom this translation of the 2wT mHealth implementation might fail or succeed [16, 17]. As part of an expansion study of the impact of 2wT on VMMC follow-up in routine settings in South Africa, we used two complementary IS frameworks to qualitatively evaluate the 2wT approach in routine practice. We applied the RE-AIM (Reach, Effectiveness, Adoption, Implementation and Maintenance) framework alongside its complement, the Practical, Robust Implementation and Sustainability Model (PRISM), to explore factors that facilitate or curtail 2wT expansion potential [18]. While the RCT and forthcoming stepped wedge study outcomes provides strong, quantitative evidence for individual (client) *reach* and *effectiveness* of RE-AIM [18], we qualitatively explored *adoption* and *implementation* at the clinician and organizational levels to enhance assessment of 2wT impact. PRISM helps explore additional contextual factors surrounding implementation including external factors that may influence 2wT maintenance.

We conducted 20 key informant interviews (KIIs) with HCWs, including clinicians, clerks, monitoring and evaluation (M&E) leads, and managers to gain insights into 2wT bottlenecks and suggestions for implementation improvements to inform next steps for 2wT expansion. We explored issues related to provider adoption (uptake) of 2wT, including soliciting perspectives on client, clinic, organization and structural factors that could influence successful scale-up. This participatory, 2wT user-centered approach also aimed to help identify technology gaps and features for improved patient, provider, and organizational experience using 2wT. Identifying further 2wT implementation enhancements could also help optimize the approach in South Africa and the region more broadly.

## Methods

### Routine VMMC service delivery

The US President’s Emergency Plan for AIDS Relief (PEPFAR) recommends VMMC for males ages 15 and older across 15 priority countries in Eastern and Southern Africa as a cost-effective components of comprehensive HIV prevention [19]. Post-VMMC follow-up at least once within 14 days is required [20], through PEPFAR specifically endorses 2wT for low-risk patients [19]. All VMMC care, regardless of follow-up method, is provided free in National Department of Health (NDoH) facilities in accordance with VMMC guidelines [21]. Clients are asked to return for scheduled day 2 and day 7 visits; some clinics or teams provide transport for clients unable to return for review. Sites used the standardized PEPFAR approach to assess, identify, and record the timing, type and severity of AEs [22]. For patients who miss day 2 visits, clinicians follow-up via phone call. VMMC teams are expected to report key PEPFAR outcomes: VMMC productivity by age group; AEs by severity and type; and # of VMMC clients with at least 1 follow-up visit within 14 days [20]. In addition to NDoH routine reporting forms, some implementing partners, including Right to Care, collect additional client M&E data from their clinician and clerical teams.

### 2wT Expansion Trial

We conducted a modified stepped wedge designed (SWD) study from January 30 – October 19, 2023, in Ekurhuleni District (Gauteng Province) and Dr. Kenneth Kaunda District (North West Province) [23]. As 2wT rolled out, clients during intervention periods were able to opt in to 2wT via Short Message Service (SMS) or choose standard of care (in-person visits). Clinicians retained discretion on 2wT enrollment; 2wT enrollment was not offered at all sites at all times. From January 30 – August 16. Between August 17 – October 19, only, WhatsApp was offered in addition to SMS as a platform for 2wT communication.

### 2wT Technology and Implementation

The 2wT system has been described in detail previously [8, 9, 24]. In brief, the 2wT model is a hybrid mHealth system that combines automated and personalized messages between clients and a VMMC nurse on SMS or WhatsApp. Males ages 15 and above with a phone present at their VMMC appointment were eligible to enroll for the opt-in 2wT follow-up approach. On the day of VMMC, clients were educated about using the 2wT follow-up option in lieu of scheduled post-operative visits and given a pamphlet with instructions and emergency contacts. After discharge, on Days 1-3, 5, 7, 10 and 13, 2wT participants were sent automated messages in the language they chose asking them to respond about complications like bleeding, swelling, or wound opening their healing. On the other days (4, 6, 8, 9, 11 and 12), they were sent educational messages. If no response was received by day 3 for minors or day 8 for adults, a nurse followed up to trace the clients and ensure healthy healing.

2wT was implemented as a hub and spoke model for the SWD study. Spokes were routine VMMC service delivery sites where non-study teams performed VMMC procedures and counseling according to NDoH standards. Spokes recruited, enrolled, and educated clients on the 2wT system and reviewed 2wT clients who needed follow-up care. The hub was a centrally located, study-specific, VMMC nurse who responded to clients, called clients who did not respond, and referred clients to spoke (facilities) for follow-up care. When clients responded to the system with a concern, the “hub nurse” responded, triaged, and referred to spoke sites if needed. Clinicians from “spoke” facilities were asked to check the 2wT system in addition to other circumcision duties. Spoke clinicians could also respond to messages when necessary. Spoke site staff were responsible for filling paper forms for routine and 2wT clients.

### Conceptual model

The conceptual model and qualitative approach was created using two common IS frameworks to guide successful translation of research to routine practice. RE-AIM [18, 25] was operationalized at the staff/setting levels (adoption and implementation) to guide measurement of what, where, when, and how the 2wT intervention was implemented (Figure 1) [26, 27]. KIIs with 2wT managers, clinicians, M&E data specialists and support staff identified insights into how to improve the 2wT system and its implementation. Second, the study team incorporated aspects of PRISM [28] to inform actionable recommendations, including internal fit (HCW and clinical context) and external fit for context (South African VMMC policies and guidelines) [29]. The combined RE-AIM and PRISM-informed model [25] (Figure 1) shaped the interview guide, analysis, and recommendations.

**Figure 1:**
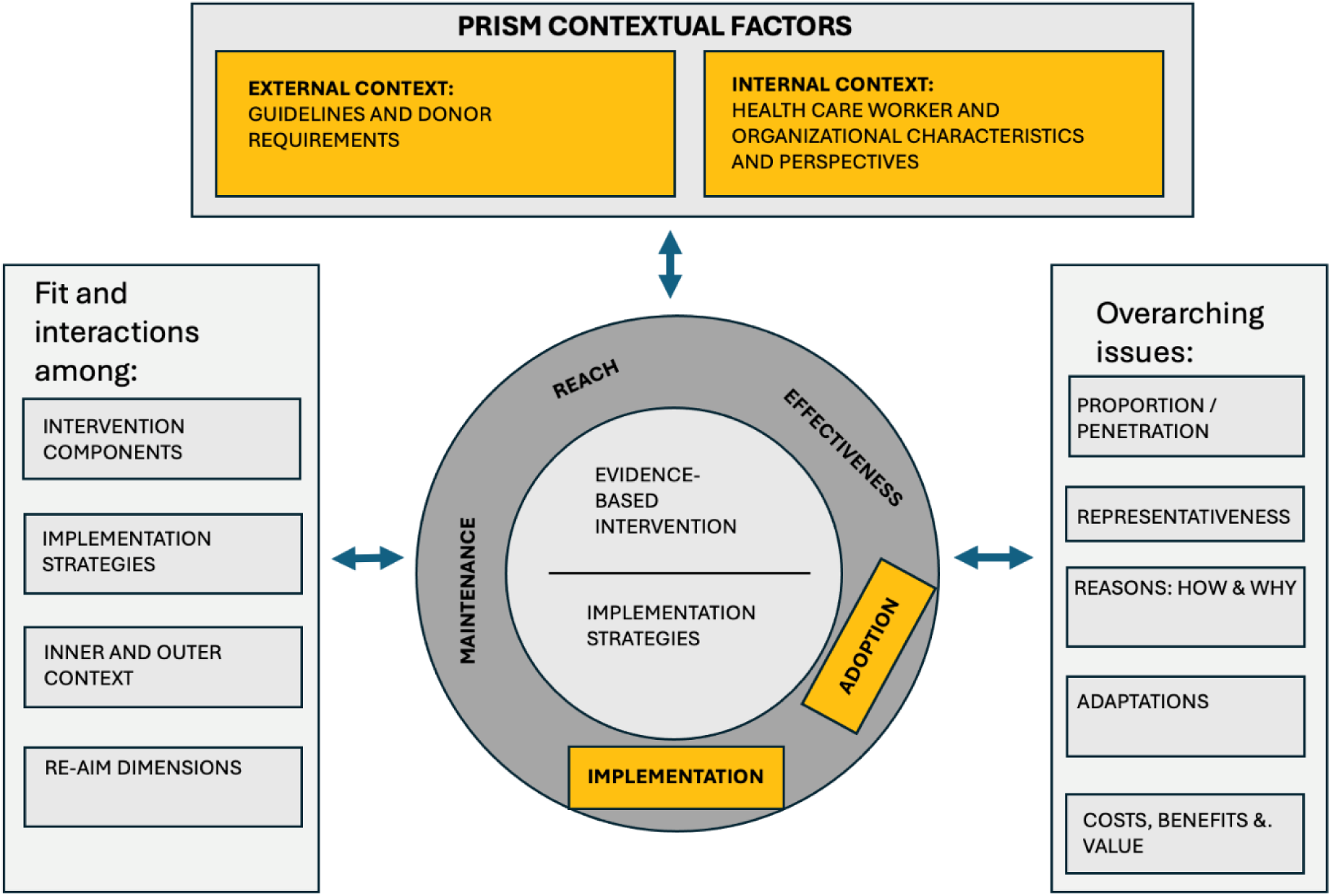
Adaptation of the Practical, Robust Implementation and Sustainability Model (PRISM) Framework* for the Stepped-Wedge Study *Model adapted from: Glasgow et al. (2019) [25] that expanded upon the PRISM framework (Feldstein & Glasgow, 2008) [28]. Constructs in yellow were explored in this study.

### Data collection and analysis

Key informant interviews (KIIs) were conducted from November 20, 2023, to January 19, 2024. Using the conceptual model, the study team developed a semi-structured interview guide, which was pilot tested through an initial practice interview in which the participant critiqued the guide and the study team made improvements. Key informants (KIs) were recruited in person from facilities involved in the expansion trial, using maximum variation sampling to include representation from those in leadership positions, clinicians, M&E data officers and demand creation officers, aiming for saturation of themes among selected respondents. A multi-lingual interviewer (author CM) conducted interviews in English; respondents also responded in IsiZulu, Sesotho or Setswana. Only the interviewer and participant were present at the time of the interview. Interviews were recorded at the participant’s respective facility and took between 12 minutes and 1 hour 23 minutes, an average of 53 minutes. Interviews were transcribed and translated into English. The interviewer and a researcher from UW analyzed interviews using rapid qualitative analysis [30, 31] and a combined deductive and inductive approach. Throughout the analytic process, the broad study team was involved in categorization, thematic analysis, and interpretation of results, raising the likelihood that interview suggestions could be translated to fit improvement. The analysts wrote 2-page summaries of each interview, created a list of themes from summaries and prepared the results using a matrix to inform recommendations. Results presented in this analysis largely exclude themes that were well explored previously.

### Positionality

The research team includes male and female individuals from UW’s I-TECH, Aurum, and Right to Care. Author CM from Aurum conducted interviews; CM is a black African female. All members of the research team were also involved in 2wT implementation and familiar with the expansion trial; however, none of the authors were primarily based at a facility implementing 2wT.

### Ethics

The review boards of UW (No. 00009703; CF) and the University of Witwatersrand, Human Research Ethics Committee (No. 200204; GS) approved the study protocol for the routine scale-up of 2wT as part of the step-wedge study. KII participants completed a written informed consent. No personal identifiers were included in the analyzed data.

## Results

Using the conceptual model, findings are presented in three thematic groupings: 1) implementation, including factors related to adoption; 2) 2wT internal program fit factors including training and staffing of clinicians and support staff; and 3) the external fit to South African context, including VMMC policies and guidelines.

### Participant Characteristics

Interviews from twenty people were included in analysis; data from one KI was excluded as they were not a direct user of the 2wT system (Table 1).

**Table 1.** Participant job categories.

| Type of Employee | Number |
| --- | --- |
| Management | 4 |
| Clinicians | 6 |
| M&E Data team | 8 |
| Support staff | 2 |
| **Total** | **20** |

### Implementation

#### Improve 2wT as a Monitoring and Evaluation (M&E) Tool

Many respondents noted advantages of 2wT for M&E. Some participants understood that 2wT improved follow-up verification, reporting and planning, including that it provides a record of patient care, improves time management and increases communication among clinicians. Other participants noted that the 2wT system allows clinicians to address tasks at their own pace and document client care within the system. They added that the system also captures patient details, so information can be accessed by teams from any location. Before 2wT, a KI reported that when males could not afford transportation to the VMMC clinic, they could be reviewed at their nearest clinic with data on local paper forms, resulting in gaps in client records. Participants also appreciated improved communication between clients and providers, remarking that clients could ask their providers questions at any time via SMS, filling in potential gaps in education. In addition, KIs thought that 2wT could improve AE reporting, since males in routine care seldom returned for their follow-up visits.

> *“Some of them would sit and not even want to come in. If they hadn’t texted via [2wT] we would not even know, because sometimes you would call a patient and they’ll say no, I’m fine. And the patient is not okay. So, when it’s a red flag on that side, then we can eventually communicate with them. So yeah, it did help a lot.” [Participant 16]*

Despite noted benefits of 2wT for M&E, many respondents noted that added burden of parallel reporting systems as clinicians are required to report all clients, including 2wT clients, on routine NDoH forms. This documentation duplication between the digital system and on paper greatly reduced enthusiasm and buy-in. One participant noted a possible solution is that 2wT could replace some of the required NDoH paper documentation.

> *“I think the amount of writing whether it’s [2wT] or follow up on paper, eventually becomes heavy on anyone. But if I was just focused on the 2-way texting, already [the provider] and the clients have typed something.” [Participant 21]*

#### Ensure 2wT clients are well educated in wound care and AE identification

Potential benefits of 2wT were muted by concerns about client safety using the telehealth approach. While some participants felt 2wT increased their ability to identify AEs, some were concerned that AEs would be missed. One participant recommended that patients first come for their 2-day check-in, and then start 2wT. Other clinicians feared that post-operative client education was not strong enough so clients would not be able to correctly identify signs of complications.

> *“Because the client is not a qualified somebody, [they will] not be able to pick it up because the client will say there is no AE. And the hub nurse would say, ‘no there is no AE’. But you find that there is an infection hiding somewhere. Only a qualified [clinician] can actually see that something is not 100 percent.” [Participant 10]*

#### Apply 2wT eligibility criteria consistently for appropriate client enrollment

Over time, providers noted that they learned to be more selective in who they enrolled into 2wT and who they required to attend in-person reviews. A few providers mentioned experiences with minors that led them to stop enrolling them or do so under more scrutiny. For example, some minors do not have a phone and use their parent or guardian’s phone, so the guardian needs to be home with the client and attentive to the messages. Also, clinicians were concerned about allowing clients to enroll via WhatsApp as clients might not have a data plan, instead requiring clients to enroll using SMS which was free. Overall, the need to consider each client, individually, and match the 2wT intervention to the client was considered prudent.

#### Enhance 2wT system functionalities

Participants recommended additional 2wT system functionalities, including offering the routine texts in more languages to better include clients from neighboring countries who speak different languages. Respondents also recommended adapting the system to allow clients to send photos or submit voice notes, reducing client literacy or language barriers. Finally, a suggestion for improved notifications was to include sound alerts when new messages came in from clients, especially for those submitting potential AEs.

### Internal fit factors

#### Offer hands-on trainings and follow-up mentoring to ensure confidence in 2wT use

Most participants found the 2wT trainings to be helpful, especially the practical elements in which trainers showed trainees how to use the system. One participant in a demand creation role (recruits and finds non-responsive clients for follow-up) noted they wanted to be more involved in trainings, *“I would like to have a proper training now … even though I’m at the field … I must be empowered” [Participant 2].* Participants preferred 2wT-specific mentoring, including in-person check-ins or virtual support through phone or video calling.

> *“We need to do some in-service training amongst ourselves, just to remind ourselves which aspects are more [important], what basis we need to touch on during the in-service training. We sometimes have weekly meetings where we sit down and talk and look at the aspects. I think those meetings can be [improved].” [Participant 8]*

#### Appoint dedicated staff for 2wT as part of routine VMMC care

Several participants mentioned that 2wT was easy to use, but comfort with the system increased over time. One expressed their slower uptake: *“We are also still new on this 2way texting. We are being resistant to change, but I think it will work better with time [Participant 19].* To speed expanded 2wT use at site-level, many participants recommended 2wT should have dedicated staff at facilities to reduce the perceived 2wT workload as additional to routine VMMC duties.

> *“If it’s just us and there are too many clients, we suffocate and there’s nothing we can do. One day we had 21 clients. [There were only two of us] and then imagine [us] having to cut and enroll. And then assuming that each client is five minutes, that’s more than an hour. Then were like, okay, let’s cut everyone then we will enroll after. … And we ended up unable to enroll the clients unfortunately, because after 6, clients had to go home.” [Participant 8]*

Moreover, respondents also thought that a dedicated staff person could help improve coordination. Monitoring clients within the 2wT system can be difficult to coordinate when in-person patients are also waiting. Learning how to balance the in-person reviews with the need to respond to 2wT clients via SMS or WhatsApp proved challenging.

> *“My sense was that it was more of us being all over and not being able to properly coordinate that, but for example, I think at some facilities you were able to see that, at least this person is the one that is doing it. Then you can see some kind of stability, but as soon as that person is not there, then there’s no one who takes over. So, there was no proper coordination of this.” [Participant 18]*

#### Identify a 2wT champion to encourage adoption of the 2wT approach

While many participants requested dedicated 2wT staff, others felt that a 2wT champion was missing to drive internal clinic enthusiasm. Some considered 2wT as an external study, not a component of routine VMMC care, with one person stating that 2wT is *“still a study, so we don’t even know if this thing works or not. But you guys are already forcing it on us.” [Participant 14]*. Another participant identified that when only one person at the clinic uses 2wT, its implementation stops when that person leaves. That same respondent noted that a champion was needed to promote the advantages of 2wT and encourage adoption at the clinic. *“We could not master…how we should be benefiting actually from 2-way texting especially because of not having the person champion the 2-way texting. [We were unable] to properly coordinate.” [Participant 18]*.

### External fit factors to context

#### Adapt VMMC guidelines to realities on the ground

Participants noted that completing all required in-person reviews was challenging. In part, many clients faced transportation issues or clients attending on Day 2 would not complete additional follow-up visits. These realities, combined with the pressure to report adherence to required visits for the funder, could lead to overreporting of post-operative visit attendance. One respondent noted they felt obligated to report near 100% follow-up even if many clients were unable or chose not to return for in-person visits.

> *“We would always have problems when we report to the funder, because the documents will not be filled. But the system will be saying 100% or 98%. If it’s not documented, it means it’s not done.” [Participant 10]*

#### Allow virtual follow up option in VMMC guidelines

Participants mentioned that enrolling clients in 2wT conflicts with the NDoH guidelines that require in-person visits, resulting in low support for 2wT uptake and expansion. *“In [District X], I know the Department of Health does not like 2wT. I don’t know if it’s because, according to the guidelines, [we] need to physically see the clients. [Participant 14]* Respondents were concerned about promoting an approach that contrasted to the in-person review requirement.

## Discussion

Conversations with KIs including clinicians, M&E data officers, VMMC managers and support staff demonstrate the complexity of moving from research to routine practice for evidence-based mHealth innovations. Overall, interviewees reported several facilitators of expansion, including that 2wT increases timely communication between patients and providers; fills potential gaps in client education; improves follow-up verification; and strengthens reporting. However, implementation was stymied by several obstacles at both provider and organizational levels, including perceptions of increased workload, lack of site champions and lack of an enabling policy environment. With competing demands on clinicians’ time and decreasing global VMMC funding, evidence-based, effective, safe, and cost-effective mHealth innovations like 2wT should be attractive at client, provider, and organizational levels. However, 2wT obstacles appear more influential than perceived benefits, reducing buy-in and momentum to scale. Application of the RE-AIM and PRISM frameworks shed light on several opportunities to energize 2wT buy-in and facilitate scaling across South Africa.

First, to create momentum for 2wT expansion, improved sensitization and dissemination efforts about 2wT benefits for clinical care are needed. During the expansion trial, the evidence of 2wT safety and efficiency benefits were not well disseminated. Without knowledge of the RCT outcomes, providers lacked confidence that 2wT could ensure quality care with referral and tracing safeguards. Moreover, 2wT awareness and education sessions for HCWs were not sufficiently prioritized, especially in light of frequent staff turnover, weakening buy-in. Additional 2wT refresher training was also needed to bolster site teams’ ability to provide 2wT-specific client education, counseling, and SMS-based encouragement. Although 2wT augments, not replaces, in-person visits, site teams needed more confidence to exercise their discretion: funneling low risk clients to 2wT and scheduling minors or clients with higher perceived AE risk to in-person reviews, streamlining follow-up care overall. With additional 2wT team engagement with site clinicians and HCWs about the implementation process, it is likely that greater buy-in could be secured from these critical stakeholders to improve the innovation [32].

Alongside efforts to promote 2wT benefits for clinicians and client care, more emphasis is needed to increase awareness of 2wT benefits for data quality and client M&E. Additional, targeted training for data officers and managers on real-time 2wT dashboards with complete client data could further build support for 2wT’s effect on data quality, garnering more manager support for the reporting benefits. At the organization level, lack of awareness of 2wT benefits for accountability, decision-making, and client verification was also evident. In this study, the hub nurse was a strong promoter of the system and drove the creation of tasks for facilities to resolve. In the future, facility teams, themselves, would need increased encouragement and supervision to complete tasks on time, trace clients and update the 2wT system. Enhanced facility staff buy-in is needed to ensure maintained 2wT implementation. To create more enthusiasm for 2wT clinical and M&E advantages, and support expansion planning, a champion from a high 2wT uptake site should be engaged in a peer-to-peer support for 2wT adoption. These champions, either hubs or spoked, could reinvigorate 2wT enthusiasm and consideration of 2wT as a locally optimized intervention [33, 34], increasing the likelihood of successful scale-up [35].

Lastly, efforts should increase focus on 2wT’s benefit to provide quality follow-up with fewer in-person reviews, rather than promote the reduction in overall VMMC workload [36]. Although 2wT decreases the number of in-person reviews and costs less in comparison to two post-operative visits, HCWs work more efficiently, but not fewer hours, using 2wT. Language matters: clear understanding of the workload implications of using 2wT are needed. Second, fewer in-person follow-up visits likely happen than reported, creating a potential false workload comparison between 2wT and routine care visits. Reported VMMC follow-up rates vary, with studies finding between 63-90% of VMMC clients attending at least one visit [8, 37–39]. However, over-reporting of follow-up visits may reflect fear of consequences, including job loss or other disciplinary measures [40]. Compared to actual post-operative visits, the time required to enroll, manage, and engage with 2wT clients may be higher. Assigning a dedicated 2wT nurse to manage telehealth clients or reallocation of team duties to incorporate 2wT activities and oversight structures into the routine workflow merits attention. As workflow concerns may reduce innovation adoption by frontline HCWs [41], identification of a site champion becomes even more critical to support expansion potential.

### Strengths and Limitations

While attention was given to maintain objectivity, the research team was involved in conducting and analyzing KIIs, a bias that cannot be quantified. Members of the NDoH were not included in KIIs; future effort would benefit from their perspective. Client perspectives [23] and common constraints of electricity, network, and funding [13, 15] were previously explored and not repeated here. Although this study aimed to enhance implantation of 2wT, suggestions to improve routine VMMC service delivery were passed to VMMC teams. The specific suggestions for 2wT features and functionality were shared with the technical team who could enhance the 2wT system at scale.

## Conclusion

Increased education, sensitization, and awareness is needed to move the 2wT approach from research to routine practice creating more enthusiasm for expansion. More attention on the clinical and M&E benefits could help create champions for 2wT scale-up, overcoming hurdles in consistent uptake. Adoption of several recommendations could push the balance in favor of expansion. First, augmented education to ensure that clients are prepared to self-monitor must be consistently implemented, ensuring that clients are informed and empowered to heal safely at home. Second, 2wT M&E processes must be streamlined, including fewer redundancies with the routine VMMC reporting alongside a more user-friendly dashboard. Third, there needs to be policy coverage to allow for the 2wT telehealth approach for clients who opt-in, allowing clinician discretion to recommend in-person reviews only when needed. With clear 2wT benefits for clients, clinicians, M&E data officers and VMMC managers, more engagement of these key stakeholders could overcome identified hurdles to 2wT expansion, creating momentum to scale.

## Author Contributions

*Conceptualization:* Caryl Feldacker, Calsile Makhele

*Formal Analysis:* Isabella Fabens, Calsile Makhele, Caryl Feldacker

*Investigation:* Calsile Makhele

*Methodology:* Caryl Feldacker, Geoffrey Setswe

*Project administration:* Caryl Feldacker, Geoffrey Setswe

*Supervision:* Nelson Igaba, Khumbulani Moyo, Felex Ndebele, Jacqueline Pienaar, Geoffrey Setswe, Caryl Feldacker

*Validation:* Calsile Makhele

*Writing – original draft:* Isabella Fabens, Caryl Feldacker

*Writing – review & editing:* Isabella Fabens, Calsile Makhele, Nelson Igaba, Felex Ndebele, Jacqueline Pienaar, Geoffrey Setswe, Caryl Feldacker

## Data Availability

All data produced in the present study are available upon reasonable request to the authors

## Appendix

## Appendix 1: Consent to Participate in KII

**PARTICIPANT INFORMATION SHEET AND INFORMED CONSENT FORMS FOR HEALTHCARE WORKERS STEP WEDGE**

**Main study**: Expanding and Scaling Two-way Texting to Reduce Unnecessary Follow-up and Improve Adverse Event Identification Among Voluntary Medical Male Circumcision Clients in Republic of South Africa

**Scale-Up:** Implementation of 2wT in Routine Practice with the Goal of Improving 2wT-based Follow-up Within the VMMC program in South Africa (**2WT-2-Scale**)

**HREC REFERENCE NO: 200204**

**PROTOCOL NO: AUR2-8-270 2WT**

**2WT STUDY**

Principal Investigators:

Caryl Feldacker, PhD, MPH, University of Washington, Seattle, WA, USA

Geoffrey Setswe DrPH, MPH, The Aurum Institute, Johannesburg

**Funding Source and/or Sponsor:**

National Institutes of Health, 9000 Rockville Pike, Bethesda, Maryland 20892, USA. PI Name: Caryl Feldacker & Geoffrey Setswe: Application ID: 1 R01 NR019229-01A1. 1-866-504-9552 (tty: 301-451-5939) or.)

**Study Contact telephone numbers:**

| Name | Position | Phone number |
| --- | --- | --- |
| **24 Hour Urgent Assistance** |  |  |
| Felex Ndebele | Project Manager | 078 149 0446 |

Good day, my name is _______________________ (INSERT NAME), I am a _______________________ (INSERT DESIGNATION) at The Aurum Institute. I would like to invite you to consider participating in an evaluation, entitled “Expanding and Scaling Two-way Texting to Reduce Unnecessary Follow-up and Improve Adverse Event Identification Among Voluntary Medical Male Circumcision Clients in Republic of South” Africa.

**What you should know about this evaluation:**

- We give you this consent so that you may read about the purpose, risks, and benefits of this evaluation.
- The main goal of this evaluation is to help VMMC clients in Ekurhuleni, Dr Kenneth Kaunda, and Dr Ruth Semotsi Mompati districts.
- We cannot promise that this evaluation will benefit you.
- We want to document your opinions about and satisfaction with, the 2-way texting (2WT texting) follow-up method
- You have the right to refuse to take part or agree to take part now and change your mind later.
- Please review this consent form carefully. Ask any questions before you make a decision.
- Your participation is voluntary.

**PURPOSE**

You are being asked to be part of an evaluation to assess the implementation of text-based follow-up after circumcision. What we learn from this evaluation will help the National Department of Health (NDOH) decide if and how to scale two-way text-based follow-up for VMMC.

Two-way texting will be implemented for approximately 800 men in all participating districts. Then, we will include up to 20 clinicians at evaluation sites, like yourself, who will also be asked questions about this evaluation and their thoughts on text-based follow-up. As a clinician, you will be asked questions such as what you know and think about the texting method.

You were asked to consider being part of this evaluation because the information you provide in this evaluation, including how satisfied you are with the texting intervention, your experience with taking care of these men via SMS, and any suggestions you have to improve the texting may improve the intervention in the future.

**PROCEDURES AND DURATION**

If you decide to participate the following procedures will happen: We will seek to meet you in a private place and seek informed consent from you. If you consent, you will go to a private space within the clinic, and we will solicit your opinions and experiences regarding deploying 2WT. We will ask you questions like, “What were the challenges of the texting system?” The interview will take approximately 30 minutes. With your consent, the interview will be recorded.

**RISKS AND DISCOMFORTS**

Taking part in the evaluation may cause some psychological discomfort because you will be asked to provide your opinions regarding text-based follow-up after surgical circumcision.

**BENEFITS and COMPENSATION**

We cannot promise that you will receive any benefits from this evaluation. The study will help the National Department of Health (NDOH) to make male circumcision SMS follow-up stronger and, potentially, more widely available. No compensation will be offered.

**CONFIDENTIALITY**

The information you give us will be kept private (in secret). All information we collected during interviews will be locked away or kept on protected computers. No one outside of the evaluation partner will know the results of the SMS input session or your interview. Any information that could be used to identify you will be shared only with your permission and will not be used in any reports from this evaluation. The recordings will be transcribed without identifiers. The voice recordings of the interviews will be destroyed one year after the activity ends. However, the link between your identifier and the transcripts will be destroyed after the records retention period required by the Aurum Institute and the University of Washington in accordance with the law. These records will be kept locked in a separate file cabinet that only evaluation staff can enter.

The Aurum Institute, Wits HREC, US Government, or University of Washington staff sometimes review studies such as this one to make sure they are being done safely and legally. If a review of this evaluation takes place, your records may be examined. The reviewers will protect your privacy. The evaluation records will not be used to put you at legal risk of harm. A description of this clinical trial will be available on http://www.clinicaltrials.gov, as required by U.S. Law. This Web site will not include information that can identify you. At most, the Web site will include a summary of the results. You can search this Web site at any time.

We have a Certificate of Confidentiality from the United States from the National Institutes of Health. These protections only apply to data held in the United States. This helps us protect your privacy. The certificate means that we do not have to give out information, documents, or samples that could identify you even if we are asked to by a court of law in the United States. We will use the Certificate to resist any demands for identifying information.

We can’t use the Certificate to withhold your research information if you give your written consent to give it to an insurer, employer, or other person. Also, you or a member of your family can share information about yourself or your part in this research if you wish.

There are some limits to this protection. We will voluntarily provide the information to:

- a member of the United States government who needs it in order to audit or evaluate the research.
- individuals at the institution(s) conducting the research, the funding agency, and other groups involved in the research, if they need the information to make sure the research is being done correctly.
- individuals who want to conduct secondary research if allowed by federal regulations and according to your consent for future research use as described in this form.
- to relevant authorities as required by other Federal, State, or local laws.

The Certificate expires when the NIH funding for this evaluation ends. Currently, this is 31 March 2025. Any data collected after expiration is not protected as described above. Data collected prior to expiration will continue to be protected

**ETHICAL APPROVAL:**

- This clinical evaluation protocol has been submitted to the University of the Witwatersrand, Human Research Ethics Committee (HREC) and written approval has been granted by that committee.
- The evaluation has been structured in accordance with the Declaration of Helsinki (last updated: October 2013), which deals with the recommendations guiding doctors in biomedical research involving human participants. A copy may be obtained from me should you wish to review it.

**FUTURE USE**

The information that we obtain from you for this evaluation might be used for future studies. We may remove anything that might identify you from the information and specimens. If we do so, that information may then be used for future research studies or given to another investigator without getting additional permission from you. It is also possible that in the future we may want to use or share evaluation information that might identify you. If we do, a review board will decide whether or not we need to get additional permission from you.

**VOLUNTARY PARTICIPATION**

It is up to you whether you want to be part of this evaluation. Your alternative is to not participate in the evaluation. If you decide to be in it, you may stop at any time. These decisions will not affect your employment or future relationship with the NDOH or its partners. If you decide to leave the evaluation, we will ask you for information about why you are choosing to leave. It is up to you whether to answer these questions.

**<u>What if you have questions about this evaluation?</u>**

You have the right to ask and receive answers to questions about this research. If you have questions, complaints, or concerns, contact the researchers listed below:

*Prof Geoffrey Setswe at 072 025 9875 or Jacqui Pienaar at 082 965 5098*

**OFFER TO ANSWER QUESTIONS**

Before you sign this form, please ask any questions on any aspect of this evaluation that is unclear to you. You may take as much time as necessary to think it over.

**AUTHORISATION**

I am making a decision about whether or not to participate in this evaluation. My signature indicates that I have read and understood the information provided above, have had all my questions answered, and I have decided to participate.

The date I sign the document to enrol in this evaluation, that is, today’s date, MUST fall between the dates indicated on the approval stamp affixed to each page. These dates indicate that this form is valid when I enrol in the evaluation but do not reflect how long I may participate in the evaluation.

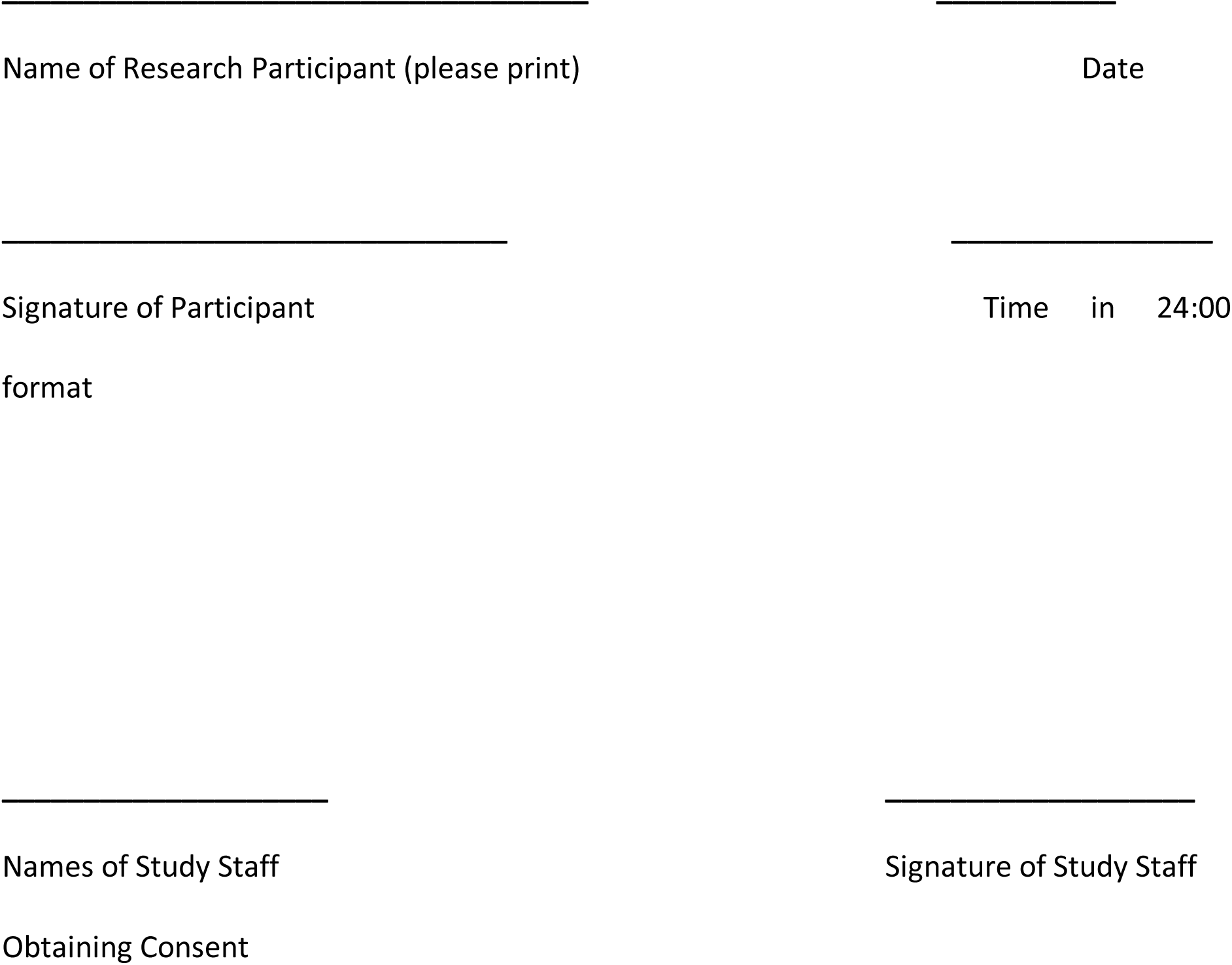
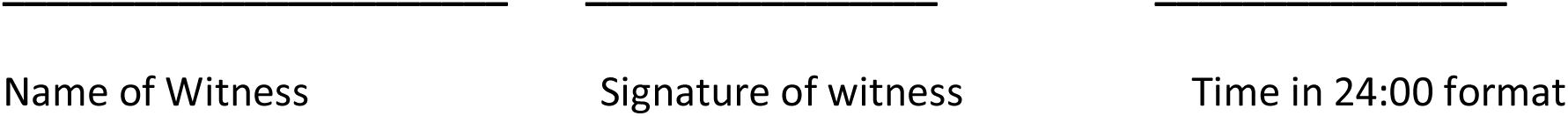

**STATEMENT OF CONSENT TO BE AUDIOTAPED**

I understand that audio recordings will be taken during the SMS input session and the evaluation interviews. *(For the statement below, please choose YES or NO by inserting your initials in the relevant box)*

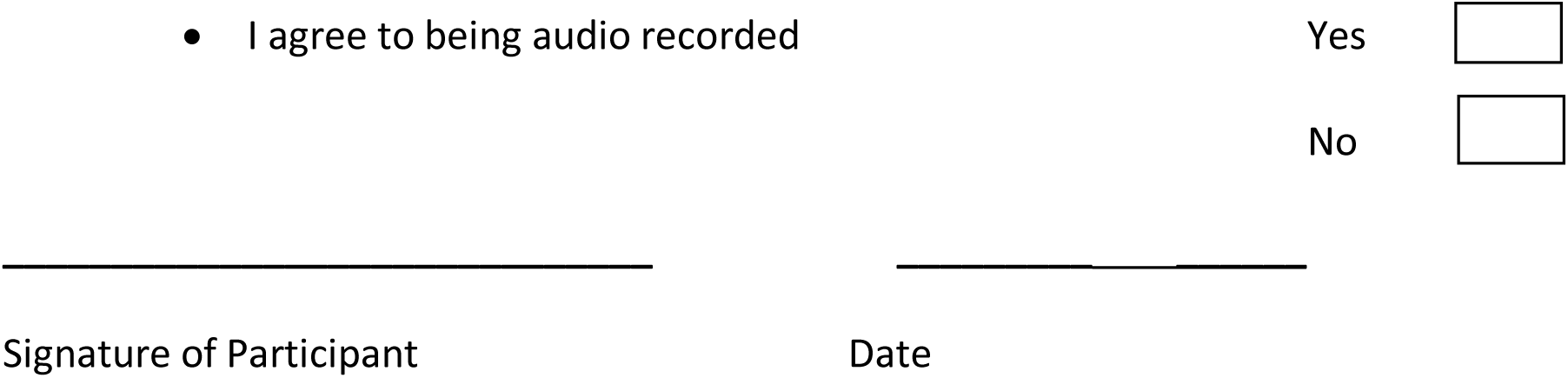

**YOU WILL BE GIVEN A COPY OF THIS CONSENT FORM TO KEEP.**

If you have any questions concerning this evaluation or consent form beyond those answered by the investigator, including questions about the research, your rights as a research subject or research related injuries; or if you feel that you have been treated unfairly and would like to talk to someone other than a member of the research team. If you want any information regarding your rights as a research participant, or complaints regarding this evaluation, you may contact Prof. Clement Penny, Chairperson of the University of the Witwatersrand, Human Research Ethics Committee (HREC), which is an independent committee established to help protect the rights of research participants at (011) 717 2301.

## Appendix 2: Key Informant Interview Guide to assess acceptability of 2WT among HCWs using CFIR framework

Adapted from: https://www.ncbi.nlm.nih.gov/pmc/articles/PMC6248861/

FIRST: Interviewer MUST verify that informed consent is signed by the interviewee

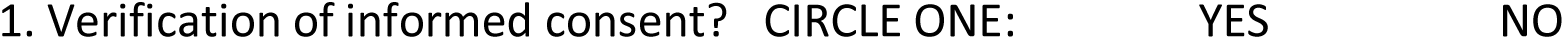

General introduction:

Hello, my name is __________________________. Thank you for agreeing to talk to me today as part of ongoing VMMC quality improvement efforts. This interview is about your experiences whilst using the two-way-texting follow-up method that your organisation has been implementing. We will explore and cover more of your thoughts, opinions, and suggestions about the texting study that you recently helped implement to inform improvements as we considering scaling up the follow-up approach. Thank you for agreeing to participate in this interview, this should take about 45 minutes of your time. I will be asking you some questions which you are free to answer in any way you wish. You may also choose not to answer any question. I encourage you to feel free to say anything concerning the topic of discussion. If a question is unclear to you, you can ask me to explain it. Your participation is voluntary and confidential. Whatever you tell me will be treated with utmost confidentiality. The information will only be used for the purposes of this program evaluation.

Please allow me to record our discussion so that I don’t miss anything. Your voice will not be heard by anyone other than our study team. Your name will not be recorded and will not appear on the transcription or any publication. The recording will be destroyed after we have prepared our transcripts.

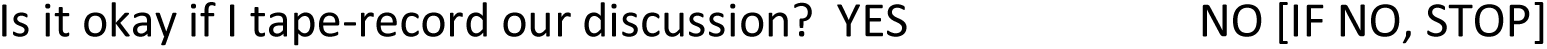

**<u>SECTION A: For all interviewees</u>**

1. To get started, I’d like to learn about your involvement with 2wT. Tell me about your role in the implementation . [**Characteristics of Individuals**] **Probe:** for all roles (site (enrolment, messaging, reviews, M&E, hub, M&E, management, oversight, reporting, etc.)

**<u>SECTION B: Only for site/hub/implementation</u>**

2. Tell me about the initial 2wT training you received. [**Inner Setting]**

a. What training materials were used to train you? **Probe/show examples:** [be specific: did they use, like, dislike, not used]: posters, flip chart, pamphlets, user guide, (toolkit of client education), HCW cheat sheets, and short videos.
b. Which one was more useful to you?
c. What were the most useful aspects of your training? **Probe:** (PowerPoint slides training, educational materials, support phone calls)
d. Tell me about refresher training sessions or on-site mentoring/support you received AFTER the program started?

- How was mentoring or support provided? **Probe:** phone, WhatsApp, Zoom? In-person? Videos?
e. How can we improve the training? **Probe:** Initial AND ongoing mentoring/support?
3. Now, let’s talk about the implementation of the 2wT approach – you may not know all aspects, but let’s talk about your experiences. From your perspective, walk me briefly through the process from client education to enrolment to interaction over 14 days to reporting. At each stage, tell me what worked well and what did not. **[Intervention Characteristics]**

a. Tell me about client sensitization on 2wT BEFORE a client is cut? [How do you introduce 2wT?]

- Tell me about post-operative counseling? Overall, how can we improve client education?
b. How about with enrolment? What works well? What does not?

- How long does the enrollment confirmation take?
- Are there differences in SMS vs. WhatsApp?
c. Regarding the Daily 2wT messaging, what worked well and what did not?
  - Following up on Potential AE reports
  - Communicating with clients
d. Regarding the Task completion, what worked well and what did not? In your opinion, how easy or difficult is it to complete tasks?

- For referrals to care
- For tracing
e. Regarding reporting and completion of the paper follow-up (review) forms, what works well and what does not? In your opinion, how easy or difficult is it to complete paper follow-up forms using information from the system?

How can we smoothen the challenges you experienced with paper and the 2wT interface? Then how can we smoothen that?

**<u>SECTION C: For all interviewees</u>**

4. Let’s talk about the effect of 2wT for VMMC at your clinic (your organization). From your perspective as a HCW, **how does 2wT impact the quality of client follow-up?**

- How does 2wT help find AEs?
  a. What is the impact of 2wT follow-up on demand creation? For HCWs, let’s talk about the impact of 2wT on your follow-up workload. How did 2wT impact your follow-up workload? [**Process**] **Probe:** How could we further improve 2wT to reduce workload?
  b. How could we improve 2wT for HCWs?
5. Tell me about how the hub nurse works with the site teams [Explain: Hub is the first line of nurse messaging and referral] **[Process of Implementation]**
  - Would you prefer the Hub nurse to be resident in your clinic or to be in a central location (one hub-to many spokes)? Why?
6. Let’s talk about scaling up. We’d like to learn about how 2wT could help more clients across more VMMC sites, in both routine and the busy season or campaign period.
  a. How many 2wT clients do you think your clinic can manage at a time?
  b. Is this with a central hub or clinic-centred hub?
  c. What influences whether this number goes up or down?
  d. How could we improve 2wT for clients?
  e. How could we improve 2wT for nurses?
  f. How could we improve 2wT for data management?
7. Let’s talk about next steps
  a. How can we make 2wT more user-friendly for workers?
  b. How can we make it more user-friendly?
  c. What new features or functions would you like to see in 2wT for MC?
  d. What are influential individuals like your immediate managers (for RtC staff) or your contractor (for GPs) saying about 2wT for VMMC? **[Process of Implementation**
  e. Where else would you like to see 2wT used **Probe**: Other healthcare contexts, locations, health areas?
8. Which opportunities do you think 2wT can bring to VMMC?
9. What are your concerns or threats you think the implementation of 2wT can bring to VMMC?
10. Before we end, do you have anything else to add for the 2wT team to improve the implementation of VMMC/2wT services?

